# Cardiorenal Outcomes with Finerenone in Patients Post-Acute Myocardial Infarction: Insights from a Global Federated Network

**DOI:** 10.64898/2026.06.24.26356503

**Authors:** Yu-Chuan Chuang, Jun-Yu Zhong, Yen-Po Lin, Tien-Chien Tsai, Hsiao-Tse Tu, Ming-Ju Chuang, Chi-Yen Wang, Wei-Wen Lin, Wen-Lieng Lee, Tsun-Jui Liu, Chung-Lieh Hung, Tze-Fan Chao

## Abstract

**Background:** Finerenone, a non-steroidal mineralocorticoid receptor antagonist (nsMRA), improves cardiorenal outcomes in chronic kidney disease, type 2 diabetes, and heart failure. However, its clinical efficacy and safety when initiated early after acute myocardial infarction (AMI) remain unknown.

**Methods:** This retrospective cohort study utilized the TriNetX global federated network to identify adult patients with AMI who initiated finerenone within 6 months of the index event. These were compared to a propensity score-matched control group of AMI survivors who did not receive finerenone.

**Results:** After 1:1 propensity score matching, 1,012 patients were included (506 per group; mean age 69 years). The cohort represented a high-risk phenotype with a high prevalence of type 2 diabetes (∼84%) and CKD (∼80%). Over a 2-year follow-up, finerenone treatment was associated with a lower risk of the composite endpoint of mortality and heart failure (HR 0.644; 95% CI 0.495–0.837; P < 0.001). Notably, finerenone was also associated with a lower risk of progression to ESRD or CKD stage 5 (HR 0.573; 95% CI 0.387–0.851; P = 0.005) and all-cause hospitalization (HR 0.602; 95% CI 0.482–0.751; P < 0.001). There was no significant difference in the risk of adverse events, including hyperkalemia, hyponatremia, and syncope or hypotension between groups.

**Conclusion:** In this real-world study of high-risk post-AMI patients, early initiation of finerenone was associated with lower risks of composite cardiovascular events, all-cause mortality, and renal disease progression, without a significant increase in adverse events. These findings warrant validation in prospective randomized controlled trials.

## Introduction

Patients after myocardial infarction (MI) remain at high risk for subsequent events, including recurrent MI, heart failure (HF), stroke, and cardiovascular death.(1) Approximately 1 in every 5 survivors of acute myocardial infarction (AMI) suffer a second cardiovascular event within the first year, despite significant advances in acute care.(2) Secondary prevention is important in patients who have experienced acute myocardial infarction, because it reduces the risk of recurrent cardiovascular events, decreases mortality, and improves long-term prognosis.(3)

Robust evidence showed that mineralocorticoid receptor antagonists (MRAs) reduces cardiovascular mortality and heart failure events in the AMI population.(4) Steroidal MRAs, both spironolactone and eplerenone, are indicated after acute myocardial infarction primarily for patients with left ventricular systolic dysfunction (LVEF≤40%) and either clinical HF or diabetes based on landmark trials such as RALES trial and EPHESUS trial.(5–7) However, for patients with acute MI without HF or reduced LVEF, the role of MRA is less clear.(8, 9) Therefore, routine use of steroidal MRAs after acute MI is recommended for patients with reduced LVEF and HF or diabetes, but not for all patients with MI.(10, 11)

Non-steroidal mineralocorticoid receptor antagonists (nsMRAs), such as finerenone, have demonstrated cardiovascular and renal benefits in patients with chronic kidney disease, type 2 diabetes, and HF.(12) Furthermore, finerenone provides benefits in recovery from myocardial infarction in animal models. In preclinical studies using rat models of MI induced by coronary artery ligation, finerenone improved left ventricular systolic and diastolic function and reduced plasma prohormone of brain natriuretic peptide levels, indicating attenuation of post-infarct cardiac dysfunction and neurohormonal activation.(13) Additional mechanistic studies in animal models demonstrate that finerenone reduces cardiac fibrosis, hypertrophy, and inflammation, as well as improves left ventricular dysfunction after MI following myocardial injury.(14, 15)

Currently, there is no direct clinical evidence to support the early use of nsMRA as alternative therapies for mineralocorticoid receptor antagonism in patients after myocardial infarction. Therefore, our aim was to conduct the study using the global federated network database to assess the patient with administration within 6 months after index AMI to investigate the potential benefits of finerenone use in the AMI population.

## Methods

### Data Source and Ethics Statement

This retrospective observational cohort study utilized the TriNetX Global Collaborative Network, a federated health research network that provides access to deidentified electronic health records (EHRs) from approximately 160 million patients in 171 healthcare organizations (HCOs) around the world. The platform harmonizes data, including demographic data (TriNetX Coding System, LOINC), diagnoses (International Classification of Diseases, Tenth Revision, Clinical Modification [ICD-10-CM]), procedures (CPT and ICD-10-PCS), medications (RxNorm, ATC), and laboratory values (TriNetX Coding System, LOINC) in the Observational Medical Outcomes Partnership (OMOP) Common Data Model. (16) Data for this retrospective cohort study were extracted and analyzed from April 1, 2026, to May 20, 2026 from the TriNetX Global Collaborative Network.

This study follows the Strengthening Reporting of Observational Studies in Epidemiology (STROBE) Reporting Guidelines. (17) As the data are deidentified and comply with the Health Insurance Portability and Responsibility Act (HIPAA), the requirement of informed consent has been waived. This study was approved by the Institution Review Board of the Taichung Veterans General Hospital (IRB approval number: CE26346A).

### Study Population

We identified adult patients (aged 18 years) with a confirmed diagnosis of AMI, including both ST-elevation myocardial infarction (STEMI) and non-ST-elevation myocardial infarction (NSTEMI), occurring on or after January 1, 2018. To mimic an “incident user” design in the post-AMI setting, patients were included in the finerenone treatment group if they had a first recorded prescription of finerenone within 6 months after index AMI diagnosis. The control group consisted of AMI survivors who did not receive finerenone within the study period (defined as 1 year prior to the index event through 3 years after the event). To minimize confounding by indication and ensure safety comparability, we excluded individuals with mechanical complications of AMI (e.g., rupture of cardiac wall, rupture of papillary muscle, hemopericardium). Patients with severe renal disease, defined as end-stage renal disease (ESRD), dependence on renal dialysis at baseline, or chronic kidney disease (CKD) stage 5, were excluded. Patients with concomitant use of strong CYP3A4 inhibitors within 1 year prior to the index event were also excluded, consistent with the medication’s safety labeling. The ICD and ATC codes used for disease and medication identification are listed in **table S1** in Supplement 1. The study eligibility criteria and baseline covariates were evaluated during the baseline period, defined as the 3-year period prior to the index event.

### Propensity Score Matching and Covariates

To account for baseline differences and potential selection bias, we performed 1:1 propensity score matching (PSM) using the TriNetX platform’s built-in greedy nearest-neighbor algorithm with a caliper of 0.1 pooled standard deviations. Covariate selection was guided by clinical relevance to cardiorenal risk. We matched for 45 variables, including demographics (age, sex, and race/ethnicity); comorbidities (hypertension, type 2 diabetes mellitus, chronic kidney disease [stages 2–4], prior HF, obesity, atrial fibrillation, cerebrovascular disease, peripheral vascular disease, chronic obstructive pulmonary disease); medications (renin-angiotensin-aldosterone system [RAAS] inhibitors [ACEi/ARBs], beta-blockers, calcium channel blockers, steroidal mineralocorticoid receptor antagonists [MRAs], HMG CoA reductase inhibitors [statins], antiplatelet agents, anticoagulants [direct factor Xa inhibitors], insulins and analogues, biguanides [metformin], glucagon-like peptide-1 [GLP-1] receptor agonists, and sodium-glucose co-transporter 2 [SGLT2] inhibitors); and laboratory values (body mass index [BMI], baseline estimated glomerular filtration rate [eGFR] calculated via the CKD-EPI 2021 equation, urine albumin-creatinine ratio [UACR], hemoglobin A1c, LDL cholesterol, N-terminal pro-B-type natriuretic peptide [NT-proBNP], and left ventricular ejection fraction [LVEF])(**Table S2**).

### Study Outcomes

The primary outcome was a cardiovascular composite defined as the time to the first occurrence of all-cause mortality, new-onset or worsening HF requiring hospitalization or urgent care. Secondary outcomes included the individual components of the primary endpoint (all-cause mortality and new-onset or worsening HF), composite renal outcome (defined as newly developed chronic kidney disease stage 5, eGFR of less than 15 mL/min/1.73m^2^, or progression to ESRD), composite outcome of sudden cardiac death, ventricular tachycardia, and ventricular fibrillation (SCD/VT/VF), ischemic stroke, and all-cause hospitalization. The safety endpoints included the incidence of hyperkalemia, hypotension or syncope, and hyponatremia. To validate the robustness of the findings and to test for residual confounders including traumatic fractures, cataract, acute appendicitis or cholecystitis, and dermatophytosis were analyzed as falsification endpoints (negative controls)(**Table S3**). Each patient was followed 28 days after the index event until the occurrence of an outcome of interest, loss of follow-up, death, administrative censoring, or a maximum follow-up period of 2 years, whichever occurred first.

### Statistical Analysis

Baseline characteristics were compared using standardized mean differences (SMD), with an SMD < 0.10 considered indicative of adequate balance. Outcomes were analyzed using Cox proportional hazards regression models to estimate hazard ratios (HR) and 95% confidence intervals (CI). The proportional hazards assumption was assessed using Schoenfeld residuals. Event-free survival probabilities were estimated using the Kaplan-Meier method, and differences between curves were evaluated using the stratified log rank test. Pre-specified subgroup analyzes were performed in key clinical phenotypes, including age (<65 versus ≧65 years), sex, ACS type (STEMI or NSTEMI), heart failure status, diabetes status, baseline eGFR strata, Renin-Angiotensin System Blockers use, Beta-blocker use at baseline, statin use and SGLT2 inhibitor use. According to the privacy policy of the TriNetX platform, the calculations for subgroups or outcomes with fewer than 10 events were suppressed to prevent potential patient re-identification and are reported as “Not Available” (N/A); therefore, these subgroups were excluded in the subgroups analysis. All statistical analyzes were performed on the TriNetX Analytics platform (TriNetX LLC, Cambridge, MA, USA) and R software (version 4.5.2; The R Foundation for Statistical Computing, Vienna, Austria). Statistical significance was defined as a two-sided P-value < 0.05.

## Results

### Patient Characteristics

We identified 147,791,523 patients with AMI on or after January 1, 2018. After exclusion of certain complications after AMI, severe renal disease, and a history of use of strong CYP3A4 inhibitors before index 1 year, 510 patients in the finerenone treatment group and 1,340,189 patients were included in the control group. After matching the propensity score, a total of 1,012 patients were included in the final analysis, with 506 patients in both group (**Figure 1**). Baseline characteristics were well balanced with no significant differences between two groups (**Figure S1**).

**Figure 1.**
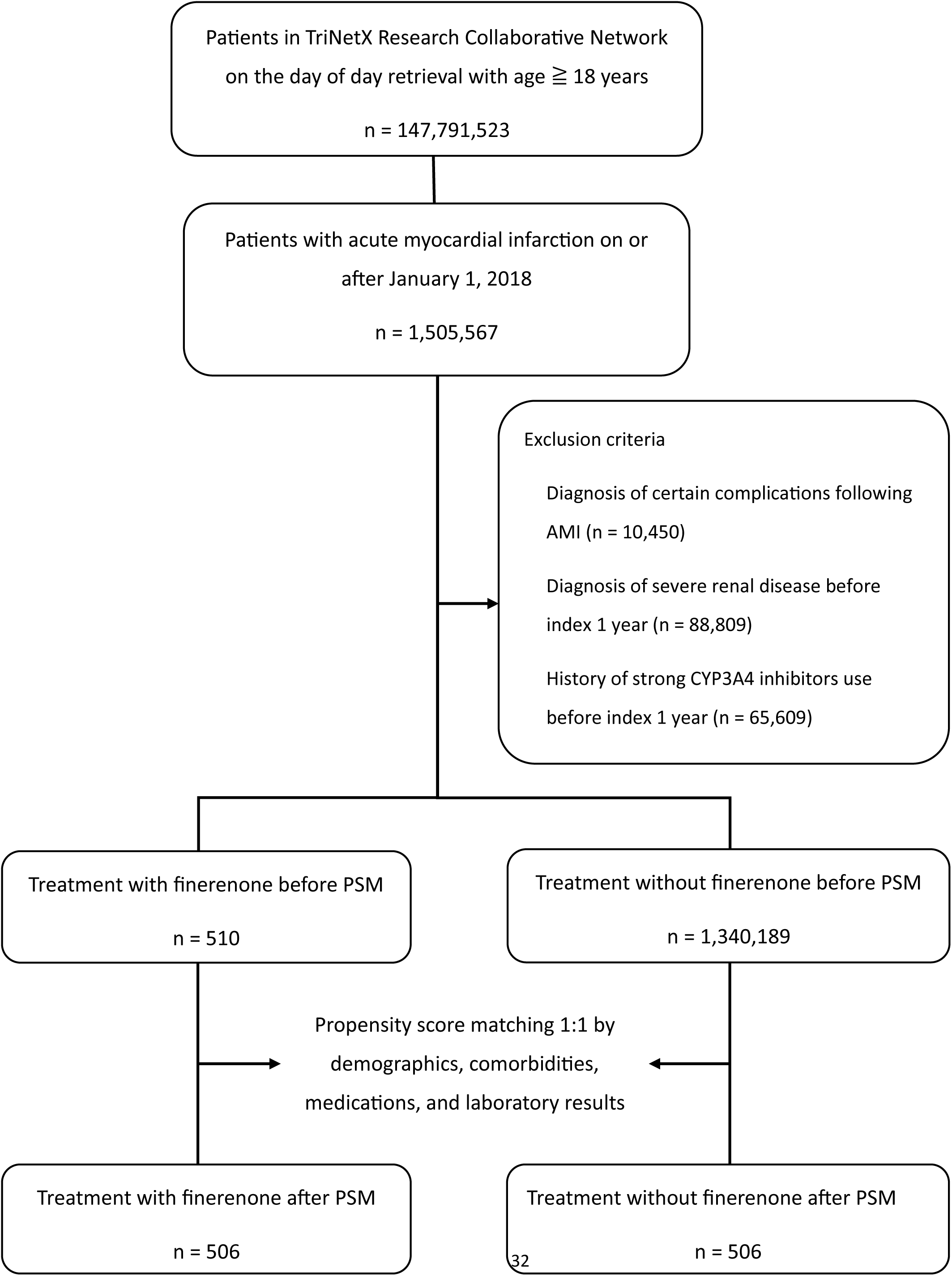
Study enrollment flowchart. The figure shows the illustration of the patient selection process for the analysis of finerenone use in patients with acute myocardial infarction. The flowchart details the exclusion criteria (post-AMI complications, severe renal disease, and CYP3A4 inhibitor use) and the sample sizes before and after propensity score matching (PSM). Abbreviations: **AMI** = acute myocardial infarction; **CYP3A4** = Cytochrome P450 3A4. Alt text: A flowchart detailing the study enrollment process, including exclusion criteria and the final sample sizes for the finerenone and control groups before and after propensity score matching.

Baseline characteristics of patients are shown in **Table 1**. The cohort was predominantly male (61.3% vs. 58.3%) and White (47.2% vs. 50.2%), with a mean age of 69.7 ± 11.1 and 69.1 ± 12.3 years, respectively. NSTEMI was the primary presentation across both cohorts (54.5% vs. 54.5%). Both groups exhibited high rates of baseline comorbidities, notably type 2 diabetes (84.0% vs. 84.2%), hypertension (81.0% in both), chronic kidney disease (78.2% vs. 81.3%), and HF (32.8% vs. 35.1%). The use of background guideline-directed medical therapies was frequent, including statins (88.9% vs. 90.9%), beta-blockers (85.1% vs. 85.4%), SGLT2 inhibitors (67.4% vs. 67.8%), and spironolactone (20.7% vs. 21.1%). Mean eGFR was 52.0 ± 24.8 versus 50.4 ± 27.5 mL/min/1.73 m², mean LVEF was 54.3 ± 12.8% versus 50.9 ± 16.6%, and mean HbA1c was uniformly 7.3% across both groups.

**Table 1.** Baseline characteristics of patients with acute myocardial infarction with or without treatment of finerenone before and after propensity score matching.

| Characteristic | Before PSM, Patient no. (%) |  |  |  | After PSM, Patient no. (%) |  |  |  |
| --- | --- | --- | --- | --- | --- | --- | --- | --- |
|  | Finerenone<br>(n = 510) | Control<br>(n = 1,340,189) | P-Value | SMD | Finerenone<br>(n = 506) | Control<br>(n = 506) | P-Value | SMD |
| **Demographics** |  |  |  |  |  |  |  |  |
| Age, mean (SD), years | 69.8 (11.1) | 67.1 (14.3) | <.001 | 0.209 | 69.7 (11.1) | 69.1 (12.3) | .421 | 0.050 |
| Male | 312 (61.2) | 794,887 (59.3) | .391 | 0.038 | 310 (61.3) | 295 (58.3) | .336 | 0.060 |
| Female | 198 (38.8) | 544,246 (40.6) | .411 | 0.036 | 196 (38.7) | 210 (41.5) | .369 | 0.056 |
| **Race** |  |  |  |  |  |  |  |  |
| White | 239 (46.8) | 794,887 (59.3) | <.001 | 0.332 | 239 (47.2) | 254 (50.2) | .345 | 0.059 |
| Black or African American | 47 (9.2) | 164,362 | .035 | 0.098 | 47 (9.2) | 49 (9.6) | .830 | 0.013 |
| Asian | 102 (20.0) | 47,600 (3.6) | <.001 | 0.527 | 98 (19.4) | 96 (19.0) | .873 | 0.010 |
| **Ethnicity** |  |  |  |  |  |  |  |  |
| Not Hispanic or Latino | 357 (70.0) | 844,762 (63.0) | .001 | 0.148 | 353 (69.8) | 360 (71.2) | .629 | 0.030 |
| Hispanic or Latino | 27 (5.2) | 48,466 (3.6) | .042 | 0.081 | 27 (5.3) | 20 (4.0) | .295 | 0.065 |
| Unknown Ethnicity | 126 (24.7) | 446,961 (33.3) | <.001 | 0.191 | 126 (25.0) | 126 (25.0) | 1 | 0 |
| **AMI type** |  |  |  |  |  |  |  |  |
| NSTEMI | 280 (54.9) | 636,778 (47.5) | <.001 | 0.148 | 276 (54.5) | 246 (54.5) | 1 | 0 |
| STEMI | 188 (36.9) | 456,441 (34.1) | .181 | 0.059 | 182 (36.0) | 169 (33.4) | .391 | 0.054 |
| **Comorbidities** |  |  |  |  |  |  |  |  |
| Type 2 diabetes mellitus | 429 (84.1) | 420,143 (31.3) | <.001 | 1.263 | 425 (84.0) | 426 (84.2) | .931 | 0.005 |
| Essential hypertension | 413 (80.9) | 805,439 (60.1) | <.001 | 0.470 | 410 (81.0) | 410 (81.0) | 1 | 0 |
| Hyperlipidemia | 378 (74.1) | 587,468 (43.8) | <.001 | 0.647 | 374 (73.9) | 385 (76.0) | .424 | 0.005 |
| Chronic kidney disease (CKD) |  |  |  |  |  |  |  |  |
| CKD, stage 2 | 53 (10.4) | 33,622 (2.5) | <.001 | 0.325 | 52 (10.3) | 67 (13.2) | .143 | 0.009 |
| CKD, stage 3a | 124 (24.3) | 46,301 (3.4) | <.001 | 0.632 | 123 (24.3) | 121 (23.9) | .883 | 0.009 |
| CKD, stage 3b | 134 (26.2) | 41,125 (3.0) | <.001 | 0.694 | 131 (25.8) | 127 (25.1) | .772 | 0.018 |
| CKD, stage 4 | 93 (18.2) | 39,186 (2.9) | <.001 | 0.513 | 90 (17.8) | 97 (19.1) | .570 | 0.035 |
| Congestive heart failure | 168 (32.9) | 205,800 (15.3) | <.001 | 0.419 | 166 (32.8) | 178 (35.1) | .425 | 0.005 |
| Atrial fibrillation and flutter | 149 (29.2) | 295,056 | <.001 | 0.165 | 149 (29.4) | 162 (32.0) | .375 | 0.055 |
| Cerebrovascular diseases | 128 (25.1) | 222,659 (16.6) | <.001 | 0.209 | 127 (25.1) | 132 (26.0) | .718 | 0.022 |
| Peripheral vascular disease | 79 (15.4) | 104,431 (7.7) | <.001 | 0.241 | 78 (15.4) | 80 (15.8) | .862 | 0.010 |
| COPD | 93 (18.2) | 209,034 (15.6) | .100 | 0.070 | 93 (18.4) | 97 (19.1) | .747 | 0.002 |
| Obesity | 143 (28.0) | 197,546 (14.7) | <.001 | 0.328 | 143 (28.2) | 151 (29.8) | .579 | 0.034 |
| **Medications** |  |  |  |  |  |  |  |  |
| ACE inhibitors | 126 (24.7) | 288,274 (21.5) | .079 | 0.075 | 126 (24.9) | 115 (22.7) | .416 | 0.051 |
| Angiotensin II inhibitor | 328 (64.3) | 241,681 (18.0) | <.001 | 1.065 | 324 (64.0) | 345 (68.1) | .163 | 0.087 |
| Sacubitril / Valsartan | 93 (18.2) | 27,097 (2.0) | <.001 | 0.557 | 91 (17.9) | 104 (20.5) | .300 | 0.065 |
| Beta blockers | 435 (85.3) | 637,129 (47.5) | <.001 | 0.872 | 431 (85.1) | 432 (85.4) | .929 | 0.005 |
| Calcium channel blockers | 348 (68.2) | 385,733 (28.7) | <.001 | 0.859 | 344 (67.9) | 329 (65.0) | .317 | 0.062 |
| Spironolactone | 107 (20.9) | 81,322 (6.0) | <.001 | 0.446 | 105 (20.7) | 107 (21.1) | .877 | 0.009 |
| Eplerenone | 13 (2.5) | 5,608 (0.4) | <.001 | 0.176 | 13 (2.5) | 16 (3.1) | .571 | 0.035 |
| HMG CoA reductase inhibitors | 453 (88.8) | 661,783 (49.3) | <.001 | 0.943 | 450 (88.9) | 460 (90.9) | .296 | 0.065 |
| Antiplatelet Medications | 409 (80.2) | 726,019 (54.1) | <.001 | 0.576 | 405 (80.0) | 401 (79.2) | .754 | 0.019 |
| Direct factor Xa inhibitors | 134 (26.3) | 137,732 | <.001 | 0.423 | 134 (26.5) | 153 (30.2) | .185 | 0.083 |
| Insulins | 273 (53.5) | 225,329 | <.001 | 0.832 | 269 (53.1) | 276 (54.5) | .658 | 0.027 |
| Biguanides | 182 (35.7) | 128,610 (9.6) | <.001 | 0.656 | 181 (35.7) | 176 (34.7) | .742 | 0.020 |
| GLP-1 analogues | 139 (27.2) | 36,815 (2.7) | <.001 | 0.730 | 136 (26.9) | 126 (24.9) | .472 | 0.045 |
| SGLT2 inhibitors | 345 (67.6) | 61,282 (4.5) | <.001 | 1.741 | 341 (67.4) | 343 (67.8) | .893 | 0.008 |
| **Laboratory and echocardiography measurements** |  |  |  |  |  |  |  |  |
| eGFR, mean (SD), mL/min/1.73m<sup>2</sup> | 51.8 (24.7) | 69.7 (27.1) | <.001 | 0.687 | 52.0 (24.8) | 50.4 (27.5) | .366 | 0.061 |
| HbA1c, mean (SD), % | 7.3 (1.7) | 6.6 (1.7) | <.001 | 0.410 | 7.3 (1.6) | 7.3 (1.8) | .624 | 0.034 |
| LDL-C, mean (SD), md/dL | 73.9 (38.7) | 93.1 (41.1) | <.001 | 0.480 | 73.9 (38.9) | 76.4 (39.1) | .389 | 0.062 |
| BMI, mean (SD), kg/m<sup>2</sup> | 29.9 (7.0) | 29.0 (7.2) | .021 | 0.121 | 29.9 (7.0) | 29.8 (7.8) | .776 | 0.020 |
| NT-proBNP, mean (SD), pg/mL | 4566.3<br>(8137.4) | 4661.9 (9134.9) | .883 | 0.011 | 4620.2<br>(8188.5) | 5172.7<br>(9233.8) | .527 | 0.063 |
| UACR, mean (SD), mg/g | 1047.2<br>(2001.6) | 386.6 (1785.7) | <.001 | 0.348 | 1055.6<br>(2027.4) | 767.5<br>(1438.4) | .154 | 0.163 |
| LVEF, mean (SD), % | 54.4 (12.7) | 53.8 (14.2) | .653 | 0.043 | 54.3 (12.8) | 50.9 (16.6) | .086 | 0.223 |

Data are presented as number (%) for categorical variables, and mean (standard deviation) for continuous laboratory and echocardiographic values.
Abbreviation: **SD**: Standard deviation, **SMD**: Standardized Mean Difference, **PSM**: Propensity score matching, **n**: Number (sample size), **AMI**: Acute myocardial infarction, **NSTEMI**: Non-ST-elevation myocardial infarction, **STEMI**: ST-elevation myocardial infarction, **CKD**: Chronic kidney disease, **COPD**: Chronic obstructive pulmonary disease, **ACE**: Angiotensin-converting enzyme, **HMG CoA**: 3-hydroxy-3-methylglutaryl-coenzyme A, **GLP-1**: Glucagon-like peptide-1, **SGLT2**: Sodium-glucose cotransporter-2, **eGFR**: Estimated glomerular filtration rate, **HbA1c**: Glycated hemoglobin, **LDL-C**: Low-density lipoprotein cholesterol, **BMI**: Body mass index, **NT-proBNP**: N-terminal pro-B-type natriuretic peptide, **UACR**: Urine albumin-to-creatinine ratio, **LVEF**: Left ventricular ejection fraction

### Primary Outcome

After 2 years of follow up, finerenone treatment was associated with a significantly lower risk of primary composite outcome compared to the control group (hazard ratio [HR], 0. 644; 95% CI, 0.495–0.837; P < .001)(**Figure 2**). A total of 87 patients (17.2%) in the finerenone group and 157 patients (31.0%) in the control group experienced the primary outcome (**Table 2** and **Figure 2A**). Regarding secondary outcomes, finerenone treatment was associated with a significantly lower risk of all-cause death (HR, 0.450; 95% CI, 0.289–0.710; P < .001)(**Figure 2B**), as well as lower risk of new-onset or worsening HF (HR, 0.730; 95% CI, 0.543–0.982; P = .0367)(**Figure 2C**). Furthermore, significant reductions were observed in the composite renal outcome (HR, 0.573; 95% CI, 0.387–0.851; P = .0051)(**Figure 2D**) and all-cause hospitalization (HR, 0.602; 95% CI, 0.482–0.751; P < 0.001)(**Figure S2**). No significant differences were observed between 2 groups regarding the risk of SCD/VT/VF or stroke (**Figure S3** and **Figure S4**).

**Figure 2.**
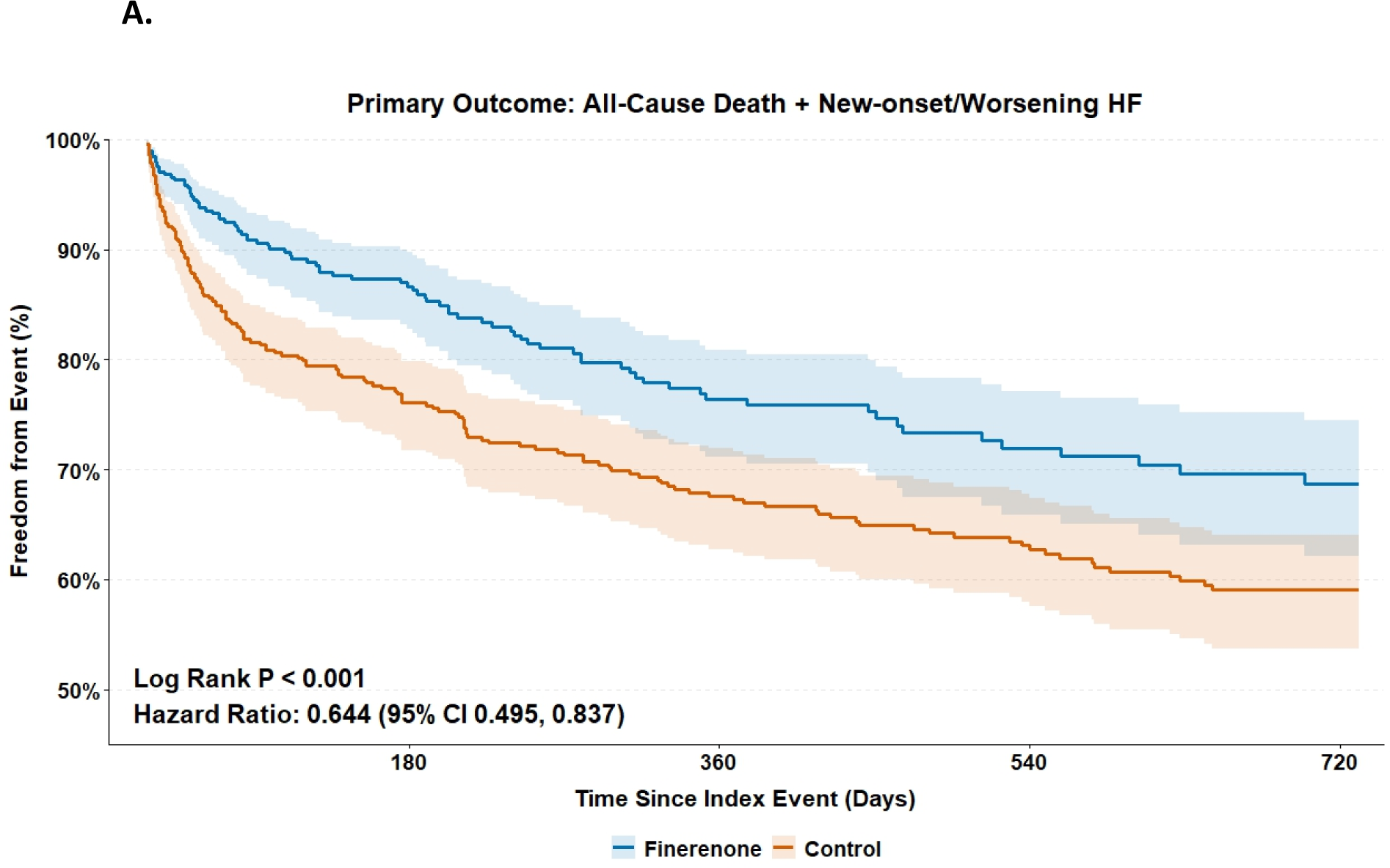
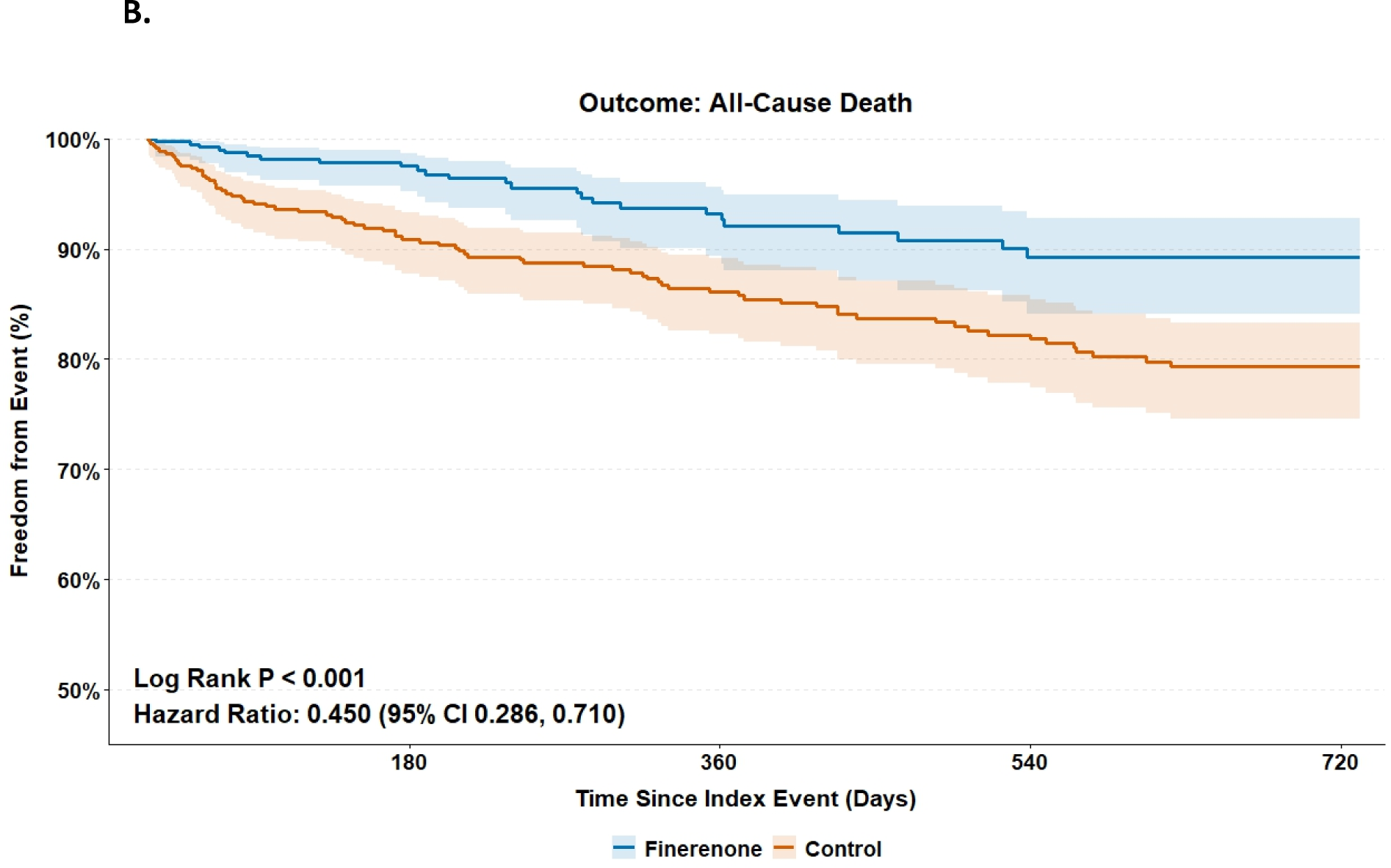
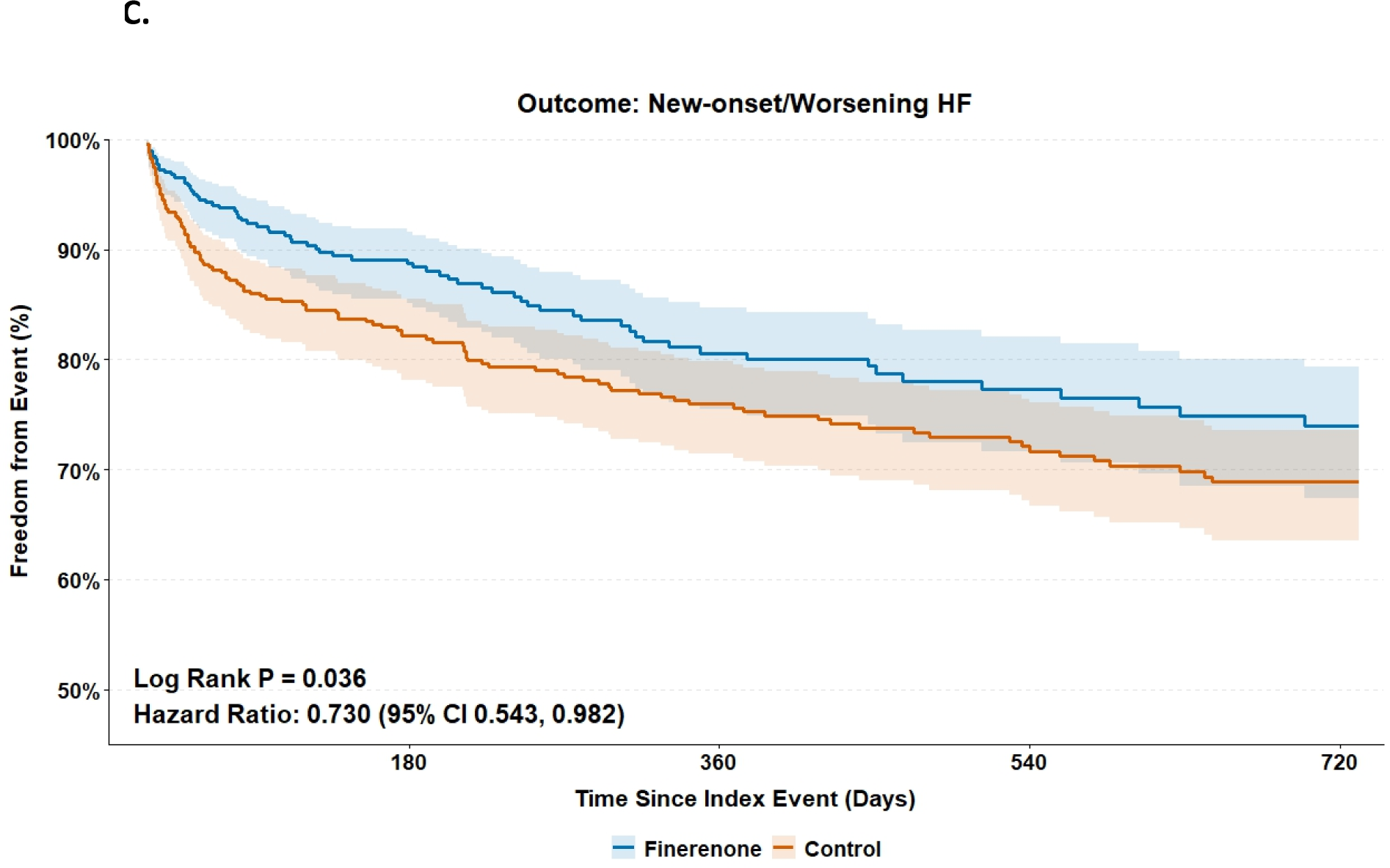
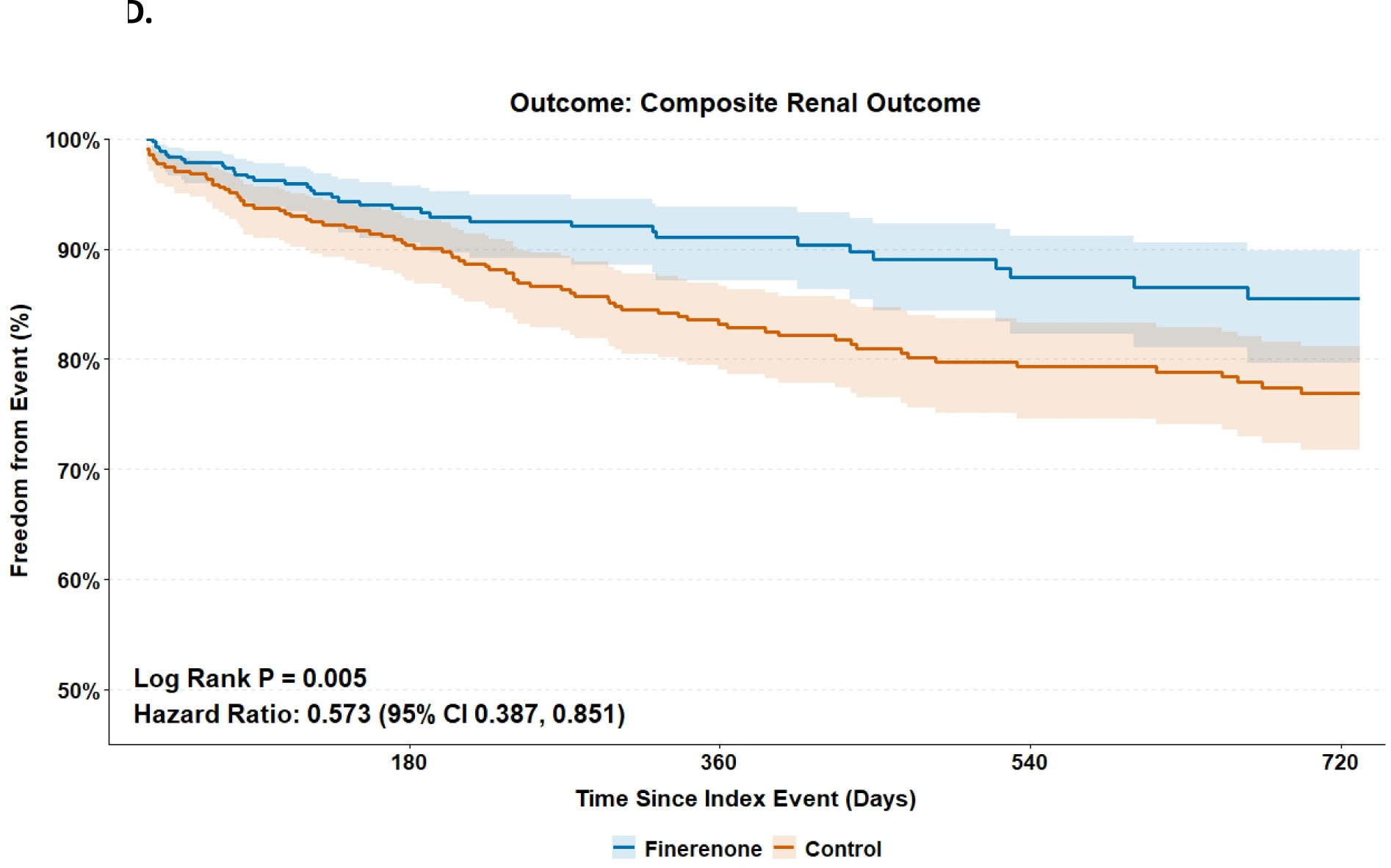
Kaplan-Meier survival curves for the primary composite cardiovascular outcome and major secondary endpoints. Finerenone was associated with a lower risk of primary composite endpoint (all-cause death and new-onset or worsening HF) (2A), all-cause mortality (2B), new or worsening heart failure (2C), and composite renal outcome (2D). Abbreviation: **HF** = heart failure Alt text: Four Kaplan-Meier survival curves demonstrating a lower risk of the primary composite cardiovascular outcome, all-cause mortality, heart failure, and composite renal outcome in the finerenone group compared to the control group over a 2-year follow-up.

**Table 2.** Primary and secondary clinical outcomes in the propensity score-matched cohort.

| Endpoint | Finerenone group<br>(n = 506) No. (%) | Control group<br>(n = 506) No. (%) | Hazard Ratio (95% CI) | Log-rank<br>P value |
| --- | --- | --- | --- | --- |
| **Primary outcome** |  |  |  |  |
| All-Cause Death and New-onset/Worsening HF | 87 (17.2) | 157 (31.0) | 0.644 (0.495, 0.837) | < .001 |
| **Secondary outcome** |  |  |  |  |
| All-Cause Death | 25 (4.9) | 74 (14.6) | 0.450 (0.286, 0.710) | < .001 |
| New-onset/Worsening HF | 71 (14.0) | 115 (22.7) | 0.730 (0.543, 0.982) | .036 |
| Composite Renal Outcome | 36 (7.1) | 80 (15.8) | 0.573 (0.387, 0.851) | .005 |
| All-Cause Hospitalization | 122 (24.1) | 222 (43.8) | 0.602 (0.482, 0.751) | < .001 |
| SCD / VT / VF | 30 (5.9) | 52 (10.2) | 0.735 (0.468, 1.155) | .179 |
| Stroke | 26 (5.1) | 52 (10.2) | 0.633 (0.395, 1.016) | .055 |

Data are presented as number (%), Hazard Ratio (HR) with 95% Confidence Interval (CI).
**Abbreviations:** HF = heart failure; SCD = sudden cardiac death; VT = ventricular tachycardia; VF = ventricular fibrillation; ESRD = end-stage renal disease.

### Subgroup Analysis

**Figure 3** presents the subgroup analyses comparing finerenone with control for the primary composite outcome. The treatment effects were generally consistent across subgroups and were similar to those observed in the overall cohort. The lower risk of the primary composite outcome associated with finerenone was particularly evident among patients younger than 65 years (HR 0.568, 95% CI 0.365–0.882), men (HR 0.597, 95% CI 0.437–0.815), those with diabetes (HR 0.604, 95% CI 0.475–0.769), advanced renal impairment (eGFR <30 mL/min/1.73 m²; HR 0.634, 95% CI 0.449–0.895), and those receiving concomitant SGLT2 inhibitors (HR 0.693, 95% CI 0.511–0.941), beta-blockers, or statins.

**Figure 3.**
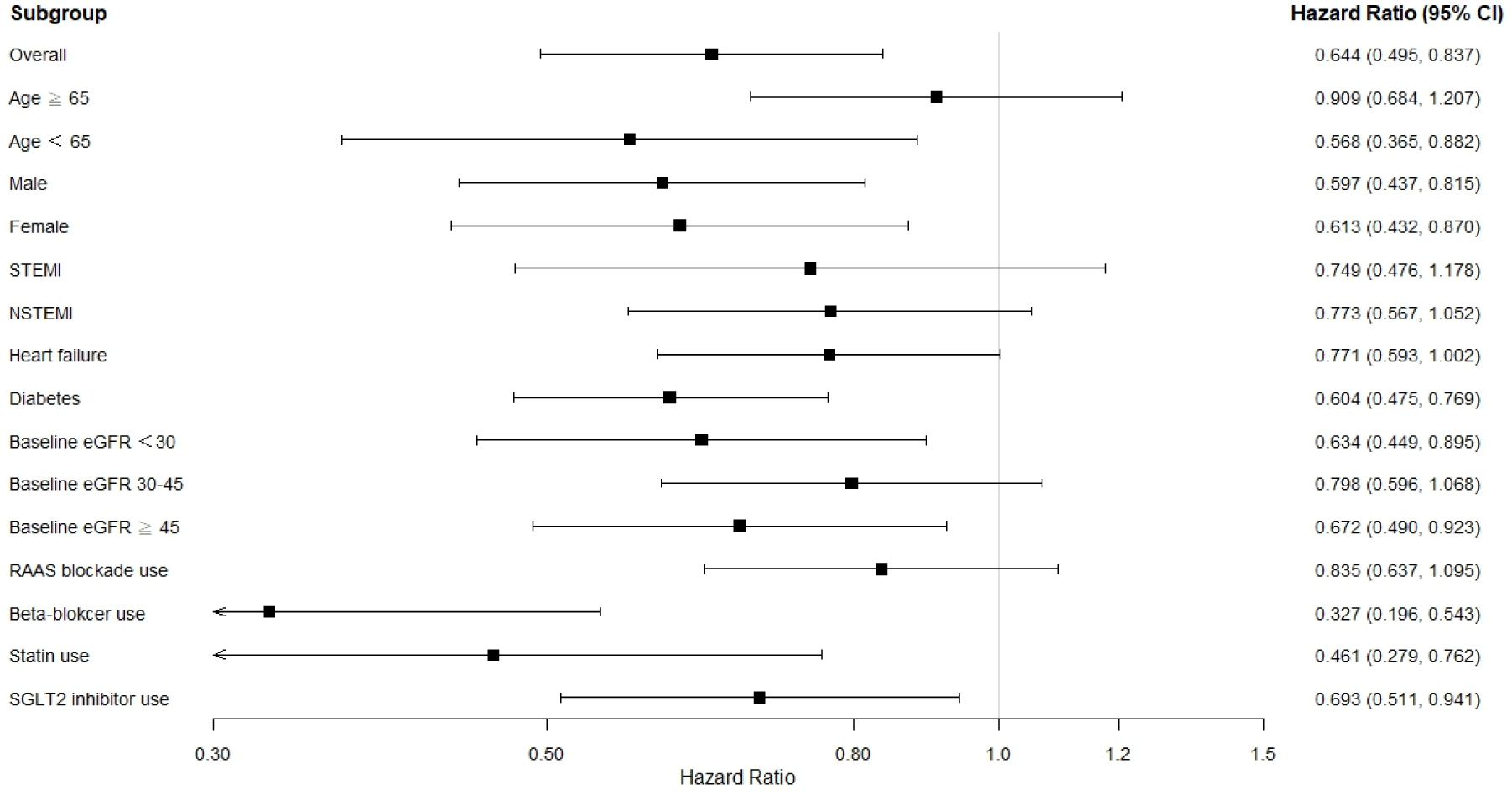
Subgroup analysis for the primary composite outcome in the propensity score-matched cohort. Subgroup analyses demonstrated consistent trends of benefit with finerenone across key clinical profiles. Significant reductions in composite cardiovascular risk were observed in patients with concomitant SGLT-2 inhibitor use, as well as across both the lowest (<30) and highest (≥45) baseline eGFR categories. The treatment effect was particularly robust and statistically significant in patients with a history of diabetes, males, females, patients under 65 years of age, and those on background therapy with beta-blockers or statins. Abbreviations: **STEMI** = ST-elevation myocardial infarction; **NSTEMI** = non-ST-elevation myocardial infarction; **eGFR** = estimated glomerular filtration rate; **RAAS** = renin-angiotensin-aldosterone system; **SGLT2** = sodium-glucose co-transporter 2 Alt text: A forest plot showing subgroup analyses for the primary composite outcome, illustrating consistent treatment benefits of finerenone across various clinical profiles, including diabetes status, eGFR categories, and concomitant medication use.

### Adverse events

There was no significant difference in the risk of hyperkalemia (9.5% in finerenone group vs. 14.0% in control group; HR, 0.893; 95% CI, 0.618–1.289; P = 0.544) or hypotension or syncope (P = 0.121) and hyponatremia (P = 0.150) between finerenone and control (**Table S4**).

### Falsification endpoints

To further assess the potential for residual confounding, we examined four falsification endpoints. No significant associations were observed between finerenone use and any of these negative control outcomes (**Table S5**). These findings suggest that the observed differences in clinical outcomes between the finerenone and control groups were less likely to be attributable to treatment selection bias or residual confounding.

## Discussion

To the best of our knowledge, this study represents the first dedicated exploration of finerenone, a novel nsMRA, in the specific context of recent AMI. In a high-risk cohort with prevalent diabetes and CKD, initiating finerenone within 6 months post-AMI was associated with a lower risk of composite cardiovascular events (HR 0.644; P < 0.001), all-cause mortality (HR 0.450; P < 0.001), new-onset or worsening HF (HR 0.730; P = 0.036) and progression to CKD stage 5 or ESRD (HR 0.573; P = 0.005).

Our study addresses a critical evidence gap in the management of acute myocardial infarction. The efficacy of steroidal MRA is well established in post-MI patients with heart failure and a reduced ejection fraction, as demonstrated by the RALES trial with spironolactone and the EPHESUS trial with eplerenone.(5, 7) However, the previous results of the ALBATROSS trial, which evaluate early spironolactone use in addition to standard therapy in post-MI patients, and recent data from the CLEAR trial both indicated no benefit from routine spironolactone use in unselected patients post-MI.(8, 9) Although previous meta-analyses of major randomized controlled trials evaluating steroidal MRAs (spironolactone and eplerenone) in post-MI populations have shown significant reductions in all-cause death (odds ratio [OR], 0.85; 95% CI, 0.76–0.95) and new or worsening heart failure (OR, 0.83; 95% CI, 0.73–0.94), the magnitude of benefit observed with early initiation of finerenone in our real-world cohort is notably greater than these historical treatment effects.(18) The benefits of finerenone in reducing new-onset or worsening HF among post-MI patients in our study align with the findings of the FIDELITY pooled analysis, which comprise the FIDELIO-DKD and FIGARO-DKD trials. This analysis demonstrated a robust 22% reduction in hospitalization for heart failure (hazard ratio [HR], 0.78; 95% CI, 0.66– 0.92) and a 23% reduction in composite kidney outcome (HR, 0.77; 95% CI, 0.67–0.88).(19–21) Consistent with our observations, the recent FINEARTS-HF trial demonstrated that finerenone significantly reduces the composite outcome of total worsening heart failure events (including hospitalizations and urgent visits) and cardiovascular death compared to placebo (rate ratio, 0.84; 95% CI, 0.74–0.95; P=0.007).(22) Additionally, the finerenone cohort demonstrated a numerically lower, albeit not significant, incidence of sudden cardiac death and malignant arrhythmias (SCD/VT/VF; HR, 0.735; 95% CI, 0.468–1.155; P=0.179). This observation aligns with the FIDELITY pooled analysis, which reported a significant reduction in sudden cardiac death with finerenone compared to placebo (HR, 0.75; 95% CI, 0.57–0.996; P=0.046) among patients with chronic kidney disease and type 2 diabetes.(23)

We hypothesize that initiating finerenone during this period of increased neurohormonal activation allows its pleiotropic effects to translate into greater absolute risk reductions than observed in lower-risk or stable populations.(24) These results extend the therapeutic scope of finerenone beyond stable diabetic kidney disease, highlighting its potential role in optimizing post-infarction care.(25) A robust mechanistic rationale supports our findings. Finerenone possesses a unique nonsteroidal structure that confers distinct pharmacokinetic and pharmacodynamic properties, including a balanced distribution between the heart and kidneys and high selectivity for the mineralocorticoid receptor (MR) without the hormonal side effects associated with steroidal agents.(26) Preclinical studies suggest that finerenone exerts more potent anti-inflammatory and antifibrotic effects than spironolactone or eplerenone by differentially recruiting transcriptional cofactors.(27) Because the acute post-MI period is driven by aldosterone-mediated maladaptive ventricular remodeling, fibrosis, and inflammation, this potent and selective MR blockade may interrupt pathological processes more effectively than delayed therapy, directly driving the substantial survival benefits observed in our study.(13, 14, 27)

Our subgroup analyses demonstrated consistent benefit across all categories of eGFR, aligning with FINEARTS-HF data in stable heart failure.(28) Our findings extend this efficacy to the vulnerable post-MI setting, providing an important therapeutic option for underrepresented patients with advanced CKD.(12, 29) Furthermore, the cardioprotective benefit was maintained regardless of the use of the SGLT2 inhibitor, consistent with recent FINEARTS-HF findings.(30) The CONFIDENCE trial substantiates the complementary mechanisms of these agents, demonstrating that combined therapy offers synergistic cardiorenal protection while mitigating the risk of hyperkalemia associated with MRA monotherapy.(31) This dual therapy approach is likely highly translatable to the high-risk post-MI population. Last but not least, the safety profile observed in our cohort is notably reassuring compared to established trial data. Our study did not find a significant excess risk; hyperkalemia rates were 9.5% in the finerenone group versus 14.0% in the control group (P = 0.544). This favorable safety signal probably reflects the distinct clinical context of the post-MI setting, where intensive electrolyte monitoring is standard, along with the high prevalence of concomitant medications such as SGLT2 inhibitors and loop diuretics, which are known to promote kaliuresis.(32, 33)

### Study limitations

This study has several limitations. First, although we employed rigorous propensity score matching to balance baseline characteristics, the potential for residual confounders of unmeasured variables, such as socioeconomic status, specific granular details of coronary anatomy, or physician preference, cannot be entirely excluded. Although the results of falsification analyses may suggest that the significant differences between finerenone and control with regard to clinical outcomes in which we were interested may be less likely due to treatment selection bias, we can only report “associations” and do not imply causality. Second, the cohort of finerenone users (n = 506) represents a small fraction of the overall AMI population screened (>1.5 million), suggesting that finerenone prescription in this setting is currently highly selective. This early adopter bias may limit the generalizability of our findings to the broader post-MI population. Third, a notable limitation is the incompleteness of the echocardiographic data; left ventricular ejection fraction (LVEF) was recorded for only 28% of the cohort. As LVEF is a primary determinant for MRA indication in current guidelines, specifically for heart failure with reduced ejection fraction (HFrEF), the lack of universal LVEF data limits our ability to strictly differentiate between guideline-directed use and novel off-label application. Fourth, the median follow-up of approximately 2 years, while sufficient to detect early separation in the Kaplan-Meier curves, may underestimate long-term benefits or late-emerging safety signals that would accrue over a decade of chronic therapy. Due to the limitations mentioned above, our findings should be viewed as hypothesis-generating and require validation in dedicated prospective randomized controlled trials.

In conclusion, our study demonstrates that finerenone initiated within 6 months of AMI is associated with significant lower risks of all-cause mortality, new-onset or worsening HF, and progression to CKD stage 5 or ESRD, without an excessive risk of adverse events. These findings suggest that finerenone may offer a valuable therapeutic strategy for this vulnerable patient population, extending its established cardiorenal benefits to the acute post-MI setting. Although promising, these observational results are hypothesis-generating and underscore the urgent need for prospective randomized controlled trials to validate this strategy.

## Data Availability

All data used for this study is publicly available by institutional agreement through the TriNetX platform. The data utilized in this study are proprietary to the TriNetX Global Collaborative Network. Under the strict data use agreements, privacy policies, and institutional agreements governing the platform, the authors do not own the data and are strictly prohibited from directly sharing, downloading, or distributing individual participant data. Qualified researchers and investigators can independently access the same datasets by establishing their own institutional agreement and credentialing with the TriNetX platform (www.trinetx.com) and executing the query parameters described in the Methods section.

## Acknowledgments

The authors did not receive any financial support for conducting this review or publication.

## Reference

1. Hall M, Smith L, Wu J, Hayward C, Batty JA, Lambert PC, et al. Health outcomes after myocardial infarction: A population study of 56 million people in England. PLoS Med. 2024;21(2):e1004343.

2. Jernberg T, Hasvold P, Henriksson M, Hjelm H, Thuresson M, Janzon M. Cardiovascular risk in post-myocardial infarction patients: nationwide real world data demonstrate the importance of a long-term perspective. Eur Heart J. 2015;36(19):1163–70.

3. Piepoli MF, Corra U, Dendale P, Frederix I, Prescott E, Schmid JP, et al. Challenges in secondary prevention after acute myocardial infarction: A call for action. Eur J Prev Cardiol. 2016;23(18):1994–2006.

4. Puglla Sanchez LR, Rojas Toledo AD, Lajczak P, Ruiz Arroyo JR. Cardiovascular effects of mineralocorticoid receptor antagonists in acute coronary syndrome: an updated systematic review and meta-analysis of randomized clinical trials. Clin Res Cardiol. 2025.

5. Pitt B, Zannad F, Remme WJ, Cody R, Castaigne A, Perez A, et al. The effect of spironolactone on morbidity and mortality in patients with severe heart failure. Randomized Aldactone Evaluation Study Investigators. N Engl J Med. 1999;341(10):709–17.

6. Alfarano M, Marchionni G, Costantino J, Ballatore F, Verardo R, Miraldi F, et al. Aldosterone-Related Cardiovascular Disease and Benefits of Mineralocorticoid Receptor Antagonists in Clinical Practice. JACC Adv. 2025;4(6 Pt 1):101762.

7. Pitt B, Remme W, Zannad F, Neaton J, Martinez F, Roniker B, et al. Eplerenone, a selective aldosterone blocker, in patients with left ventricular dysfunction after myocardial infarction. N Engl J Med. 2003;348(14):1309–21.

8. Beygui F, Cayla G, Roule V, Roubille F, Delarche N, Silvain J, et al. Early Aldosterone Blockade in Acute Myocardial Infarction: The ALBATROSS Randomized Clinical Trial. J Am Coll Cardiol. 2016;67(16):1917–27.

9. Jolly SS, d’Entremont MA, Pitt B, Lee SF, Mian R, Tyrwhitt J, et al. Routine Spironolactone in Acute Myocardial Infarction. N Engl J Med. 2025;392(7):643–52.

10. Byrne RA, Rossello X, Coughlan JJ, Barbato E, Berry C, Chieffo A, et al. 2023 ESC Guidelines for the management of acute coronary syndromes. Eur Heart J. 2023;44(38):3720–826.

11. Rao SV, O’Donoghue ML, Ruel M, Rab T, Tamis-Holland JE, Alexander JH, et al. 2025 ACC/AHA/ACEP/NAEMSP/SCAI Guideline for the Management of Patients With Acute Coronary Syndromes: A Report of the American College of Cardiology/American Heart Association Joint Committee on Clinical Practice Guidelines. J Am Coll Cardiol. 2025;85(22):2135–237.

12. Harrington JL, Canonico ME, El Rafei A, Solomon SD, Teerlink JR, Vaduganathan M, et al. Nonsteroidal and Steroidal Mineralocorticoid Antagonists: Rationale, Evidence, and Unanswered Questions. JACC Heart Fail. 2025;13(10):102637.

13. Kolkhof P, Delbeck M, Kretschmer A, Steinke W, Hartmann E, Barfacker L, et al. Finerenone, a novel selective nonsteroidal mineralocorticoid receptor antagonist protects from rat cardiorenal injury. J Cardiovasc Pharmacol. 2014;64(1):69–78.

14. Lavall D, Jacobs N, Mahfoud F, Kolkhof P, Bohm M, Laufs U. The non-steroidal mineralocorticoid receptor antagonist finerenone prevents cardiac fibrotic remodeling. Biochem Pharmacol. 2019;168:173–83.

15. Gueret A, Harouki N, Favre J, Galmiche G, Nicol L, Henry JP, et al. Vascular Smooth Muscle Mineralocorticoid Receptor Contributes to Coronary and Left Ventricular Dysfunction After Myocardial Infarction. Hypertension. 2016;67(4):717–23.

16. Palchuk MB, London JW, Perez-Rey D, Drebert ZJ, Winer-Jones JP, Thompson CN, et al. A global federated real-world data and analytics platform for research. JAMIA Open. 2023;6(2):ooad035.

17. Cuschieri S. The STROBE guidelines. Saudi J Anaesth. 2019;13(Suppl 1):S31–S4.

18. d’Entremont MA, Cheema Z, Pitt B, Kedev S, Cornel JH, Stankovic G, et al. Mineralocorticoid Receptor Antagonists in Myocardial Infarction Patients: A Systematic Review and Meta-Analysis of Randomized Trials. JACC Heart Fail. 2025;13(9):102531.

19. Agarwal R, Filippatos G, Pitt B, Anker SD, Rossing P, Joseph A, et al. Cardiovascular and kidney outcomes with finerenone in patients with type 2 diabetes and chronic kidney disease: the FIDELITY pooled analysis. Eur Heart J. 2022;43(6):474–84.

20. Pitt B, Filippatos G, Agarwal R, Anker SD, Bakris GL, Rossing P, et al. Cardiovascular Events with Finerenone in Kidney Disease and Type 2 Diabetes. N Engl J Med. 2021;385(24):2252–63.

21. Bakris GL, Agarwal R, Anker SD, Pitt B, Ruilope LM, Rossing P, et al. Effect of Finerenone on Chronic Kidney Disease Outcomes in Type 2 Diabetes. N Engl J Med. 2020;383(23):2219–29.

22. Solomon SD, McMurray JJV, Vaduganathan M, Claggett B, Jhund PS, Desai AS, et al. Finerenone in Heart Failure with Mildly Reduced or Preserved Ejection Fraction. N Engl J Med. 2024;391(16):1475–85.

23. Filippatos G, Anker SD, August P, Coats AJS, Januzzi JL, Mankovsky B, et al. Finerenone and effects on mortality in chronic kidney disease and type 2 diabetes: a FIDELITY analysis. Eur Heart J Cardiovasc Pharmacother. 2023;9(2):183–91.

24. Galuppo P, Bauersachs J. Mineralocorticoid receptor activation in myocardial infarction and failure: recent advances. Eur J Clin Invest. 2012;42(10):1112–20.

25. Filippatos G, Kapelios CJ. Mineralocorticoid Receptor Antagonists in Post-Myocardial Infarction Patients: An Old Relationship Revisited? JACC Heart Fail. 2025;13(9):102521.

26. Agarwal R, Kolkhof P, Bakris G, Bauersachs J, Haller H, Wada T, et al. Steroidal and non-steroidal mineralocorticoid receptor antagonists in cardiorenal medicine. Eur Heart J. 2021;42(2):152–61.

27. Kolkhof P, Lawatscheck R, Filippatos G, Bakris GL. Nonsteroidal Mineralocorticoid Receptor Antagonism by Finerenone-Translational Aspects and Clinical Perspectives across Multiple Organ Systems. Int J Mol Sci. 2022;23(16).

28. Ostrominski JW, Mc Causland FR, Claggett BL, Desai AS, Jhund PS, Lam CSP, et al. Finerenone Across the Spectrum of Kidney Risk in Heart Failure: The FINEARTS-HF Trial. JACC Heart Fail. 2025:102439.

29. Lv R, Xu L, Che L, Liu S, Wang Y, Dong B. Cardiovascular-renal protective effect and molecular mechanism of finerenone in type 2 diabetic mellitus. Front Endocrinol (Lausanne). 2023;14:1125693.

30. Vaduganathan M, Claggett BL, Kulac IJ, Miao ZM, Desai AS, Jhund PS, et al. Effects of the Nonsteroidal MRA Finerenone With and Without Concomitant SGLT2 Inhibitor Use in Heart Failure. Circulation. 2025;151(2):149–58.

31. Agarwal R, Green JB, Heerspink HJL, Mann JFE, McGill JB, Mottl AK, et al. Finerenone with Empagliflozin in Chronic Kidney Disease and Type 2 Diabetes. N Engl J Med. 2025;393(6):533–43.

32. Fioretti F, Testani JM, Tio MC, Pitt B, Butler J. Aldosterone and Aldosterone Modulation in Cardio-Kidney Diseases. J Am Coll Cardiol. 2025;86(5):354–73.

33. Morales J, Handelsman Y. Cardiovascular Outcomes in Patients With Diabetes and Kidney Disease: JACC Review Topic of the Week. J Am Coll Cardiol. 2023;82(2):161–70.

